# Assessing Equity and Representativeness in Randomised Controlled Trials: A Feasibility Study

**DOI:** 10.64898/2026.06.25.26356548

**Authors:** Chizoba Oparah, Hannah O’Keefe, Opeyemi Agbeleye, Jane Nesworthy, Gill Norman, Tafadzwa Patience Kunonga

**Affiliations:** NIHR Innovation Observatory, Newcastle University, Newcastle-upon-Tyne, United Kingdom; Population Health Sciences Institute, Newcastle University, Newcastle-upon-Tyne, United Kingdom

**Keywords:** Randomised Controlled Trials, Horizon scanning, representativeness, equity, PROGRESS-Plus framework

## Abstract

Clinical trials often enrol populations that differ from those who ultimately receive the interventions, raising concerns about external validity and health equity. Trial registries could provide an early opportunity to assess representativeness, but it is unclear whether registry data contain sufficient information to enable such assessments. This study evaluated the feasibility of using registry data to assess representativeness in Phase II and III pharmacological randomised controlled trials. A search of ClinicalTrials.gov (December 2024 to January 2025) identified trials with results posted after 1 January 2023 across cardiovascular disease (CVD) excluding stroke, diabetes mellitus, and selected mental health disorders. Of 1,328 records screened, 98 trials met inclusion criteria (51 Phase III, 47 Phase II). Reporting completeness was variable, particularly in Phase II studies. CVD and diabetes trials predominantly included middle-aged to older adults, while mental health trials recruited mainly individuals aged 36 to 50 years. Across CVD and mental health trials, participants were largely male. Reporting of BMI, contraception, and comorbidity criteria was inconsistent, though available data suggested these factors influenced sample composition. Fewer than 10% of trials reported equity-relevant characteristics beyond age and sex, and none addressed intersectionality. Assessing equity using registry data is feasible but constrained by incomplete and inconsistent reporting.

## Introduction

Randomised controlled trials (RCTs) are widely regarded as the most robust method for evaluating the efficacy and safety of health interventions. However, strong internal validity does not guarantee that trial findings are applicable to the populations who will receive these interventions in routine practice.^1^ A longstanding concern is that many RCTs enrol participant samples that differ from the wider patient population. These differences often relate to characteristics such as age, sex, and racial or ethnic background, which can limit generalisability and reduce the usefulness of evidence for decision-making.^2,3^

One way of conceptualising this challenge is through proportional representativeness. This refers to the extent to which the composition of a trial population reflects, in broadly similar proportions, the characteristics of the population the trial is intended to inform.^3^ In principle, representativeness can be assessed by comparing participant characteristics with an appropriate reference population. In practice, assessing proportional representativeness is often constrained by incomplete or inconsistent reporting of participant characteristics. Additional uncertainty arises around which population should be used as the appropriate reference.

These limitations have implications for equity. Previous research has shown that clinical trials frequently underrepresent population groups who experience disproportionate disease burden, including women, older adults, people with multimorbidity, and individuals from socioeconomically disadvantaged or ethnically minoritised backgrounds.^2-4^ Such patterns may arise through restrictive eligibility criteria, recruitment practices, or structural barriers to participation. As a result, treatments may be evaluated in populations that do not fully reflect those most affected by disease, raising concerns about the equity and applicability of the resulting evidence.^5^

If trial evidence disproportionately reflects groups that are easier to recruit or retain, it may be less informative for groups who already experience barriers to care or worse health outcomes. This does not imply that lack of proportional representativeness directly produces inequitable outcomes, but it does increase the risk that evidence used in policy and practice will have uneven relevance. Reflecting this concern, research funders and regulators increasingly emphasise inclusive research design. In the UK, organisations such as the National Institute for Health and Care Research (NIHR) highlight the importance of generating evidence that is relevant to populations served.^6^

Despite this policy focus, there is no routine or standardised approach to assessing proportional representativeness early in the evidence lifecycle. Trial registries represent a potential source of information because they provide structured, publicly available data on trial design, eligibility criteria, recruitment plans, and, in some cases, participant characteristics (ClinicalTrials.gov, n.d.).^7^ However, registries are not designed specifically for assessing representativeness, and the extent to which they capture equity-relevant characteristics remains uncertain.^8,9^ This raises a practical feasibility question about whether trial registries contain sufficient and consistent information to support even a basic assessment of proportional representativeness across trials.

To address this question, we use the PROGRESS-Plus framework as a structured way to identify whether registry records report participant characteristics commonly associated with differences in health opportunities and outcomes, including, Place of residence, Race/ethnicity/culture/language, Occupation, Gender/sex, Religion, Education, Socioeconomic status, Social capital, plus additional factors such as age, disability, or comorbidity (Cochrane Equity Methods Group).^10^ Applying this framework enables a systematic assessment of whether trial registries provide sufficient information to examine equity-relevant dimensions of representativeness.

### Research Aim and objectives

#### Aim

To assess whether trial registry data allow identification of equity-relevant characteristics and preliminary assessment of proportional representativeness in RCTs.

#### Objectives

1. To identify which equity-relevant participant characteristics, as defined by the PROGRESS-Plus framework, are reported in trial registry records,
2. To examine the consistency and completeness of reporting of these characteristics across trials.
3. To explore whether registry-reported data are sufficient to support preliminary approaches to assessing proportional representativeness across trials.

## Methods

This study used a registry-based secondary analysis to assess the feasibility of identifying equity- relevant participant characteristics and evaluating proportional representativeness in RCTs. The analysis was structured using the PROGRESS-Plus framework, which captures characteristics associated with differences in health opportunities and outcomes.^10^ The study protocol was registered on the Open Science Framework.^11^

This study focused on pharmacological Phase II and III RCTs addressing three high-burden conditions: cardiovascular disease (CVD), diabetes mellitus, and selected mental health disorders. These conditions were chosen because they represent major contributors to morbidity and healthcare demand and are frequently prioritised in research ^12^ and policy agendas. An overview of the characteristics of these clinical areas is provided in Table 1.

### Eligibility Criteria

Studies were included if they were randomised controlled trials (RCTs) of pharmacological interventions with parallel group allocation, registered on ClinicalTrials.gov, and reporting protocols and results accessible through trial records or journal articles. Included trials focused on CVDs, diabetes, or mental health conditions, involved adult participants (≥18 years), and were classified as Phase II or III. Participant demographic characteristics were extracted when reported; where these data were unavailable, they were recorded as “not reported”. Terminated trials were included where an associated publication was available. Non-pharmacological interventions, observational studies, pilot or feasibility studies were excluded.

Mental health conditions cover a broad spectrum of indications. When used as a generic search term, this results in over 10,000 clinical trial records on clinicaltrials.gov. Therefore, we have limited to mental health conditions categorised in the UK as ‘common mental disorders’ (depression and/or anxiety, obsessive compulsive disorder (OCD), panic disorder, and stress), and well documented indications categorised as ‘other mental disorders’ ^13^(post-traumatic stress disorder (PTSD), bipolar, and schizophrenia). Full details of the inclusion criteria are available in Table 2.

### Data Sources and Search Strategy

The search strategy was designed in collaboration with an experienced information specialist (HOK). Searches were run in ClinicalTrials.gov on 9th December 2024 and designed using the conditions/disease field for the following concepts: CVD and diabetes. Searches for mental health conditions were run on the 9th of January 2025, again using the conditions/disease field, and included: depression and/or anxiety; post-traumatic stress disorder (PTSD); obsessive compulsive disorder (OCD); panic disorder; stress; bipolar; and schizophrenia. A date limitation was applied for all records with results first posted on or after 1/1/2023.

### Screening and Study Selection

All identified citations were collated and imported into EndNote 21.^14^ The deduplicated set of records was subsequently imported into Covidence for further review. ^15^A structured three-step screening process was employed to ensure methodological rigour and consistency. First, a piloting phase was conducted in which all three researchers (CO, OA and JN) collaboratively screened a random sample of 10 records from each clinical area to refine the eligibility criteria; discrepancies were resolved by consensus. In the second phase of the title and brief description screening, one researcher independently screened all remaining records, with 20% assessed in parallel by a second researcher for consistency. Records with uncertainties were assessed by a second researcher, and any unresolved cases were discussed until consensus was reached. At the third phase, full publications and associated protocols were retrieved by an information specialist (HOK), a random sample across the clinical areas was piloted by the three researchers to refine eligibility criteria. The remaining records were screened against inclusion criteria by one researcher, with 20% checked independently by another. Discrepancies across all phases were resolved through discussion or, when needed, by a third researcher.

### Data Extraction

Following the selection of studies, to ensure rigour and consistency, the data extraction process mirrored the study selection process. Using Microsoft Excel, data extraction was piloted, and refined collaboratively by the researchers (CO, OA and JN). One researcher initially extracted data from a random 10% subset of included studies, which was independently verified by a second researcher to ensure alignment on the extraction fields. Discrepancies identified during this phase were resolved through discussion, with necessary adjustments made to the extraction form. Subsequently, one researcher extracted data from the remaining studies, with a second researcher independently cross-checking 20% for accuracy. Any disagreements were resolved through discussion, with a third researcher consulted when needed. The data extraction process involved two key stages: Planned population characteristics and Actual enrolled participant data. From the trial records we extracted variables including eligibility criteria (inclusion and exclusion), and key trial characteristics such as funding source and trial phase. Demographic data for enrolled participants, including age distribution, sex/gender, ethnicity, and geographic locations, were extracted from trial results or associated publications. Adopting the PROGRESS-Plus framework, data such as age, sex, ethnicity, comorbidities, language proficiency, and other eligibility restrictions were extracted from the inclusion and exclusion criteria of each trial’s registry entry and/or protocol and published results to enable systematic identification of equity-relevant indicators and potential sources of exclusion across both planned and actual trial populations, respectively.

### Data Analysis and Synthesis

We systematically mapped the reporting of eligibility and exclusion criteria in trial registries to the PROGRESS-Plus framework and additional context-specific factors to identify equity-relevant indicators and potential sources of exclusion. For each PROGRESS-Plus domain, we applied a binary coding approach, recording whether information was explicitly reported or not. Coding decisions followed the PROGRESS-Plus framework; for example, language ability was coded within the Race/Ethnicity domain. Instances where factors were mentioned indirectly or only in passing were also recorded as “reported” if they provided sufficient information to infer the characteristic. For enrolled participant characteristics, we extracted demographic variables: age, sex/gender, race/ethnicity, location, and patient recruitment methods where these were available from registry entries or linked publications. Due to substantial variability in reporting quality across trials, the analysis focused primarily on the availability and completeness of these data, rather than attempting to quantify representativeness. This approach allowed us to assess the feasibility of using registry data to capture equity-relevant information and to identify gaps in reporting across ongoing RCTs. Results were narratively synthesised using exploratory analysis to identify equity-related patterns and comparative analysis to examine similarities and differences across studies. We also employed descriptive analysis to summarise key trial characteristics, providing an overview of the implications for equity in trial design.

## Results

A systematic search of the trial registry identified a total of 1,375 studies. Forty-seven duplicate records removed, leaving 1,328 unique studies for title and abstract screening. Of these, 987 studies were excluded, and the remaining 341 studies were assessed for full-text eligibility. A final set of 98 studies met the inclusion criteria and were included in the analysis (Figure 1).

### Characteristics of Included Studies

A total of 98 clinical trials were included in the analysis, comprising 51 Phase III and 47 Phase II trials. The trials spanned a study initiation period from 2005 to 2022, with anticipated or actual completion dates ranging from 2015 to 2027. Of the included trials, 88 had been completed, two were ongoing with posted results and eight had been terminated at time of this study. We have outlined the characteristics of included phase II and III trials in Table 3.

Cardiovascular disease was the most frequently studied condition in both phases, representing nearly half of Phase II trials (46.8%) and over two fifths of Phase III (41.2%), while diabetes and mental health conditions accounted for the remaining distribution with similar proportions across phases. Trial settings were notable for substantial reporting gaps, particularly in Phase II, where two thirds of studies did not specify a setting. Among those reporting, outpatient clinics and academic or research centres featured more prominently in Phase III (27.5% and 21.6%) than in Phase II (10.6% and 12.8%).

Funding patterns showed a clear predominance of private or industry support, especially in Phase II (74.5% vs. 55.0% in Phase III), while public funding contributed to a smaller subset of trials. Recruitment performance was also mixed: 27.5% of phase III and 19.1% of phase II reported exceeding enrolment targets while 21.6% of phase III and 25.5% of phase II did not meet their recruitment target. However, more than half of all trials in both phases did not report recruitment outcomes and planned recruitment durations.

Accessibility features varied considerably across trials. The majority of studies reported the requirement for participants to travel to trial sites (Phase III: 58.9%; Phase II: 61.7%), while far fewer provided reimbursement for associated costs (Phase III: 15.7%; Phase II: 17.0%) or offered language support (Phase III: 11.8%; Phase II: 2.1%). Digital access requirements were common (Phase III: 41.2%; Phase II: 44.7%), underscoring the growing reliance on technology enabled participation.^16^

### Equity-Relevant factors Reported in the Eligibility Criteria

Reporting of equity relevant inclusion criteria varied across trials as shown in Table 4. Phase III studies most frequently reported gender/sex (74.5%), followed by BMI thresholds and time related relationship criteria. In Phase II studies, age was the most commonly reported inclusion criterion (97.9%), alongside gender/sex (72.3%). Other PROGRESS Plus characteristics, including socioeconomic status (SES), religion, social capital, and place of residence, were rarely reported. More than 80% of trials did not provide information on education, occupation, living situation, or related social determinants.

Exclusion criteria showed a similar pattern, with most trials reporting only a limited set of PROGRESS Plus characteristics. Pregnancy was the most commonly reported exclusion across both phases (Phase II; 65.9% and Phase III; 64.7%), followed by fertility related requirements. Only isolated trials reported exclusions based on disability, age thresholds, or relationship characteristics. No trial reported exclusions based on SES, education, religion, occupation, or place of residence, and none included intersectional exclusions involving combinations of PROGRESS Plus domains.

### Patterns in eligibility criteria and participant composition

Where sufficient data were available, trial eligibility criteria and reported participant characteristics were examined to explore patterns in sample composition across clinical areas. Due to substantial variability in reporting, these analyses focused on age, gender/sex, BMI criteria, contraception requirements, and comorbidity exclusions, which were the most consistently reported variables.

Across CVD trials, age eligibility was generally broad, yet enrolled participants were predominantly older adults (mean age approximately 71.44 years), and cohorts were largely male dominated (37 trials). BMI and contraception-related eligibility criteria were reported in a minority of studies, and although fertility-related restrictions were noted, available data were insufficient to determine their overall influence on the sex distribution of participants. In diabetes trials, both sexes were consistently reported with only a modest male predominance. No trial applied sex-based exclusion criteria. Age eligibility was broad in most studies, resulting in cohorts largely composed of middle aged and older adults (40-58, 59-75) with limited inclusion of younger (<40 years) or very old (>75 years) participants. Those without BMI limits enrolled slightly smaller cohorts but substantially more women; and comorbidity exclusions were associated with smaller sample sizes and lower female representation. Mental health trials similarly permitted wide age and sex eligibility but enrolled predominantly male participants, typically in early to mid-adulthood (ranged 36 to 50 years) with insufficient reporting on BMI or contraception-related criteria to determine their effects. Overall, eligibility criteria varied across clinical areas and were associated with differences in cohort size and gender distribution, although many trial records lacked sufficient detail to determine the specific influence of individual eligibility criteria on participant enrolment. Summary of this reporting is shown in Table 5.

## Discussion

### Summary of main findings

This feasibility study examined equity-relevant reporting across 98 Phase II and Phase III clinical trials in CVD, diabetes, and mental health, using the PROGRESS-Plus framework as an analytical lens. Few trials reported PROGRESS Plus characteristics beyond age and sex, restricting insight into representativeness across place, race/ethnicity, education, occupation, socioeconomic status, or social capital. This reporting gap limits equity informed appraisal of external validity.^17 10^

Despite broad sex eligibility in most trials, CVD trials recorded persistent sex imbalance, cohorts remained male dominant, consistent with prior evidence of systematic under enrolment of women in CVD trials.^18^ Similarly with age, studies were concentrated in older adulthood for CVD trials and early to mid-adulthood for the mental health trials. Reporting on contraception and BMI criteria were also limited for the mental health and CVD trials. In diabetes, trials without BMI thresholds tended to recruit slightly fewer number of participants with higher female participation. It is generally shown that women tend to have higher rates of obesity than males.^19^ Many studies have argued that BMI is not a reflective measure of body composition. Instead, age and sex adjustments are required to reflect level of adiposity.^20^ Although obesity is not always reflected through BMI, this restriction may still influence female participant enrolment in trials. This is particularly interesting when considering the role of obesity in the development of diabetes, whereby males are more likely to develop diabetes at a younger age with lower BMI, thus BMI restrictions are likely to be less influential on male enrolment.^21^ Diabetes trials with explicit comorbidity exclusions tended to recruit slightly lower cohorts with lower female participation. Research has indicated that women have higher rates of comorbidity than males which might go some way towards explaining this.^22,23^ Furthermore, comorbidities are considered as the presence of two or more co-occurring conditions. This means that someone having a disability or frailty alongside another condition may be seen as having comorbidities, thus excluding them from enrolling.^24^ Nuances such as these must be considered during the study design phase so that what constitutes a comorbidity is clearly defined.

Pregnancy and fertility-related requirements constituted the most common exclusion criteria across both trial phases. Exclusions related to pregnancy/contraception, comorbidities, and thresholds such as BMI, disproportionately burden women of reproductive age, particularly those with caring responsibilities or lower socioeconomic resources.^25^ Digital access requirements in approximately 44% of studies, with minimal accompanying language or financial support, raise further concerns about the structural exclusion of older adults, those with lower digital literacy, and populations in lower-income or rural settings .^26^ While direct influence cannot be established from reporting alone, these patterns align with longstanding concerns that restrictive or poorly justified criteria can narrow participation and reduce generalisability.^27^

Our findings that equity relevant data are rarely reported in clinical trials aligns with studies showing that the inclusion of socioeconomic, demographic, and geographical factors (such as the PROGRESS- Plus framework) in trial publications is limited, inconsistent, and often overlooked.^28,29^ These disparities have been linked to factors including restrictive eligibility criteria, recruitment practices, and structural barriers to participation. Our findings reinforce the rationale underlying recent reporting initiatives such as Consolidated Standards of Reporting Trials (CONSORT Equity)^30^, which aims to improve transparency in reporting equity-relevant factors in clinical trials. Updated reporting standards, including CONSORT 2025^31^, emphasise the importance of clearly documenting participant characteristics and eligibility criteria to enable evaluation of the populations to whom trial findings may apply. Our study suggests that current registry reporting practices remain insufficient to feasibly support such assessments at scale. Adopting these standards ensures that RCT evidence can be effectively used to identify and address health inequalities.

### Implications for research, practice, and policy

The findings highlight several implications for improving equity in clinical research. First, trial registries could play an important role in enabling early assessment of representativeness if equity-relevant characteristics were reported more consistently. Embedding structured reporting fields aligned with frameworks such as PROGRESS-Plus could help standardise the reporting of participant characteristics and eligibility criteria.

Second, greater attention is needed at the trial design stage. Protocols should clearly specify equity-relevant eligibility and recruitment considerations and provide explicit justification for eligibility criteria that may disproportionately restrict participation, including fertility or contraception requirements, BMI thresholds, and comorbidity exclusions. Reporting standards such as SPIRIT (Standard Protocol Items: Recommendations for Interventional Trials) and CONSORT-Equity ^30,32^provide guidance for improving transparency in protocol development and trial reporting. Similarly, the NIHR INCLUDE framework^6^, emphasises the importance of designing trials that proactively address barriers to participation among underrepresented groups.

Finally, improving the reporting of equity-relevant characteristics would support more robust monitoring of representativeness in clinical research. Policymakers and health system decision-makers increasingly rely on clinical trial evidence to inform regulatory approval, reimbursement decisions, and guideline development. Strengthening the reporting of participant characteristics could enable more systematic comparisons between trial populations and the populations affected by disease, particularly if trial registry data were linked with population health datasets or disease registries.

### Strengths and limitations

This study provides a feasibility-focused analysis across a relatively large sample of Phase II and Phase III trials spanning multiple disease areas. The use of structured search strategies, duplicate screening, and independent data verification strengthened the methodological rigour of the analysis. In addition, applying the PROGRESS-Plus framework allowed a systematic assessment of equity-relevant reporting across trials.

However, several limitations should be considered. First, the analysis relied on information reported in trial registries and associated publications, which varied substantially in completeness and quality. As a result, some equity-relevant characteristics may have been considered in trial design but not reported in registry records. Second, the study focused on a limited number of clinical areas and pharmacological trials, which may limit generalisability to other types of interventions. Finally, due to incomplete reporting of participant characteristics, it was not possible to conduct formal assessments of proportional representativeness or intersectional analysis.

## Conclusion

This feasibility study demonstrates that identifying equity-relevant participant characteristics from Phase II and Phase III clinical trial registries is possible but substantially constrained by inconsistent and incomplete reporting. Across cardiovascular disease, diabetes, and mental health trials, registry records rarely reported PROGRESS-Plus characteristics beyond age and sex, limiting the ability to assess proportional representativeness or examine intersectional dimensions of inequality.

These findings highlight important operational limitations in current trial registry reporting and suggest that trial registries alone are currently insufficient to support systematic assessment of representativeness. Strengthening the reporting of equity-relevant participant characteristics through initiatives such as CONSORT-Equity, SPIRIT guidance, and the NIHR INCLUDE framework, may improve the transparency and applicability of clinical trial evidence for diverse populations.

Improving the completeness and consistency of equity-relevant reporting in trial registries could support more robust evaluation of whether clinical trials generate evidence that is relevant to the populations most affected by disease.

## Supporting information

Figure 1: Identification of studies via databases and registers

Table 1: General characteristics of each clinical area

Table 2: Eligibility Criteria

Table 3: Characteristics of included phase II and III trials

Table 4: Summary of Equity-Relevant Reporting in Eligibility Criteria

Table 5: Summary of Eligibility Criteria and Their Influence on Sample Composition trials in all clinic areas

## Data Availability

All data relevant to the study are included in the article or uploaded as supplementary information.

## Statements and Declarations

### Author Contributions

GN and TPK conceptualised the study. CO, OA, and JN conducted data curation and writing of the original manuscript draft. HO sourced the data, verified the accuracy of the data analysis and ensured adherence to reporting standards. TPK and GN provided supervision and oversaw project administration. All authors contributed to critical revision of the manuscript and approved the final version for publication.

### Ethical approval

Ethical approval was not required because this study used publicly available, aggregate secondary data from trial registries and linked publications, and did not involve collection of new data from human participants.

## Acknowledgements

We are grateful to the NIHR Innovation Observatory Public Advisory Group (PAG) for their interest in this project and their support and advice. We consulted and engaged with the PAG which brings together people from diverse backgrounds and experiences to help shape our research. For this project, the PAG provided us with insight into some key factors influencing trial representativeness. Their insights helped identify important details to consider, ensuring our research is as inclusive and meaningful as possible.

