## Supplementary material for "Assessing Equity and Representativeness in Randomised Controlled Trials: A Feasibility Study": Figure 1: Identification of studies via databases and registers

Records removed *before screening*:

Duplicate records removed

(n = 47)

Records identified from*:

Databases (n = 1375)

**Identification**

Records screened

(n = 1328)

Records excluded**

(n = 987)

Reports sought for retrieval

(n = 341)

Reports not retrieved

(n = 0)

**Screening**

Reports assessed for eligibility

(n = 341)

Reports excluded:

Wrong location (n = 23)

Wrong intervention (n = 18)

Wrong study design (n= 129)

Wrong patient population (n = 73)

Studies included in review

(n = 98)

**Included**
