## Supplementary material for "Assessing Equity and Representativeness in Randomised Controlled Trials: A Feasibility Study": Table 1: General characteristics of each clinical area

| **Clinical Area** | **Burden and Demography** | **Non-Modifiable Factors** | **Modifiable Factors** | **Inequalities and Disparities** |
| --- | --- | --- | --- | --- |
| **Cardiovascular Disease (CVD)** | Leading global cause of death, particularly among adults younger than 65; affects men and women; disproportionately impacts disadvantaged & ethnically diverse groups.^33,34^ | Age, sex, family history, ethnicity. | Smoking, poor diet, inactivity, obesity, hypertension, high cholesterol, diabetes.^35^ | Higher prevalence in socioeconomically deprived groups; approximately 20.4% of people with CVD or respiratory illness live in poverty.^36^ |
| **Diabetes (T1D and T2D)** | A chronic metabolic disease: T1D typically has a younger onset and an autoimmune basis while T2D commonly has an adult onset, linked to obesity and sedentary lifestyle; more common in certain ethnic groups.^37,38^ ^39^ | Age, ^40,41^sex (T1D is higher in males, T2D occurs more in females),^42^ethnicity: (higher in African American, Hispanic/Latino), family history. | Obesity, sedentary lifestyle, poor diet, comorbidities (hypertension, dyslipidaemia, obesity).^43^ | Higher prevalence in lower socioeconomic groups; strong link with obesity and inactivity. |
| **Mental Health** | Affects nearly 1 billion globally; one in four adults England annually; 8 in 100 people weekly with common disorders.^44^ ^45,46^ | Age, sex, genetic predisposition, disability. | Social determinants (poverty, discrimination, isolation), stigma, poor care access. | Impacts across PROGRESS-Plus domains (gender, age, SES, disability, minority groups); under-resourced systems; higher premature mortality (10–20 year's loss for severe conditions).^44^ |
