## Supplementary material for "Assessing Equity and Representativeness in Randomised Controlled Trials: A Feasibility Study": Table 2: Eligibility Criteria

| **Criterion** | **Inclusion Criteria** | **Exclusion Criteria** |
| --- | --- | --- |
| **Population** | Male and female adults (≥18 years) with cardiovascular disease (CVD), diabetes, or mental health conditions | Paediatric participants (<18 years) |
| **Intervention** | Pharmacological interventions | Non-pharmacological interventions (e.g., surgery, medical devices) |
| **Comparator** | All comparators | No comparator |
| **Outcomes** | All outcomes related to CVD, diabetes, or mental health conditions as reported in the studies | Outcomes related to other conditions |
| **Study Design** | Randomised clinical trials (parallel group allocation), Phase II or III; Registered on ClinicalTrials.gov; Published results including articles, protocols, or terminated trials with publications; Trials conducted in countries recognised by UK reference regulators; Published in English | Non-randomised or comparative studies; Phase I/II trials; Observational, pilot, or feasibility studies; Not published in English.  Trials conducted in countries NOT recognised by UK’s reference regulators. ^47^ |
| **Data Reporting** | Adult participants with demographic data (age, ethnicity, race, etc.) | Paediatric participants: Trials not reporting demographic data |
| **Measures** | Trials reporting planned trial design, recruitment plans, and actual recruitment outcomes | Trials not reporting trial design, recruitment plans, or recruitment outcomes |
