## Supplementary material for "Assessing Equity and Representativeness in Randomised Controlled Trials: A Feasibility Study": Table 3: Characteristics of included phase II and III trials

| **Characteristics** | **Phase II (n = 47)** | **Phase III (n = 51)** |
| --- | --- | --- |
| **Disease area** |  |  |
| Cardiovascular disease | 22 (46.8%) | 21 (41.2%) |
| Diabetes | 11 (23.4%) | 16 (31.4%) |
| Mental health | 14 (29.8%) | 14 (27.5%) |
| **Trial setting** |  |  |
| Hospital | 5 (10.6%) | 7 (13.7%) |
| Outpatient clinic | 5 (10.6%) | 14 (27.5%) |
| Academic/research centres | 6 (12.8%) | 11 (21.6%) |
| Investigator site | – | 4 (7.8%) |
| Community/field | – | 1 (1.9%) |
| Not reported | 31 (66.0%) | 14 (27.5%) |
| **Funding source** |  |  |
| Private/industry | 35 (74.5%) | 28 (55.0%) |
| Public | 12 (25.5%) | 9 (17.6%) |
| Not reported | – | 14 (27.4%) |
| **Recruitment success** |  |  |
| Exceeded target | 9 (19.1%) | 14 (27.5%) |
| Did not meet target | 12 (25.5%) | 11 (21.6%) |
| Not reported | 26 (55.3%) | 26 (51.0%) |
| Planned recruitment period reported | 4 (8.5%) | 5 (9.8%) |
| **Accessibility measures** |  |  |
| Travel required | 29 (61.7%) | 30 (58.9%) |
| Reimbursement offered | 8 (17.0%) | 8 (15.7%) |
| Language support | 1 (2.1%) | 6 (11.8%) |
| Digital access required | 21 (44.7%) | 21 (41.2%) |
