## Supplementary material for "Assessing Equity and Representativeness in Randomised Controlled Trials: A Feasibility Study": Table 4: Summary of Equity-Relevant Reporting in Eligibility Criteria

| **Equity-Relevant Reporting in Eligibility Criteria** | | | | |
| --- | --- | --- | --- | --- |
| **PROGRESS Plus factor** | **Inclusion Criteria** | | **Exclusion Criteria** | |
|  | **Phase III (n=51)** | **Phase II (n=47)** | **Phase III (n=51)** | **Phase II (n=47)** |
| Place of residence (e.g. rural/urban) | 2 (3.9%) | 1 (2.1%) | 0 | 0 |
| Race/Ethnicity (e.g., ethnic group, language fluency) | 6 (11.8%) | 3 (6.3%) | 0 | 0 |
| Occupation (e.g., employment status, profession-specific inclusion) | 3 (5.9%) | 1 (2.1%) | 0 | 0 |
| Gender/sex (e.g., women, men) | 38 (74.5%) | 34 (72.3%) | 0 | 0 |
| Religion (e.g., faith-based criteria) | 0 | 0 | 0 | 0 |
| Education (e.g., minimum academic level) | 7 (13.7%) | 2 (4.3%) | 0 | 0 |
| Socioeconomic status (e.g., income) | 0 | 0 | 0 | 0 |
| Social capital (e.g., community networks) | 0 | 0 | 0 | 0 |
| **Plus factors (e.g., additional considerations not covered under core PROGRESS, such as:)** |  |  |  |  |
| Disability | 3 (5.9%) | 0 | 1 (1.9%) | 4 (8.5%) |
| Age | 6 (11.8%) | 46 (97.9%) | 2 (4.0%) | 0 |
| Pregnancy | 0 | 0 | 33 (64.7%) | 31 (65.9%) |
| BMI | 15 (29.4%) | 14 (29.8%) | 1 (1.9%) | 0 |
| Fertility status/Contraception | 7 (13.7%) | 23 (48.9%) | 7 (13.7%) | 15 (31.9%) |
| Features of relationships | 3 (5.9%) | 3 (1.5%) | 8 (15.7%) | 0 |
| Time-dependent relationships | 8 (15.7%) | 2 (4.3%) | 1 (1.9%) | 11 (23.4%) |
