## Supplementary material for "Assessing Equity and Representativeness in Randomised Controlled Trials: A Feasibility Study": Table 5: Summary of Eligibility Criteria and Their Influence on Sample Composition trials in all clinic areas

| **Clinical Area** | **Domain** | **Criterion / Category** | **Trials (n)** | **Overall Pattern / Interpretation** |
| --- | --- | --- | --- | --- |
| **CVD** | **Age** | Broad (18–99 years) | 36 | Broad eligibility showed majority of the participants (n=38,113); however, reported ages clustered around older adulthood across trials. |
|  |  | Restricted (50–90 years) | 7 | Narrower criteria yielded smaller number of participants (n=2,816) but similar mean age profiles due to the older age focus of CVD trial populations. |
|  | **Gender / Sex** | Open to all sexes (no sex-based exclusions) | 43 (Male dominant in 37 trials) | women were enrolled in smaller numbers than men, resulting in predominantly male cohorts despite broad sex eligibility criteria. |
|  | **BMI Criteria** | BMI based exclusions applied | 5 | BMI thresholds (18–45 kg/m²) were used in a minority of studies; reporting did not allow determination of their effect on enrolment. |
|  |  | No BMI criteria | 38 | Most trials did not apply BMI restrictions. |
|  | **Contraception Requirements** | Contraception or fertility related requirements | 13 | Requirements for highly effective contraception were common, but reporting was insufficient to assess their influence on cohort and gender composition. |
|  |  | Not reported / not required | 30 | Limited reporting prevented assessment of impact on enrolment. |
| **Diabetes** | **Age** | Broad (18–99) | 22 | Broad age limits captured most participants (n=10,778), with cohorts dominated by middle-aged and older adults. |
|  |  | Restricted (18–65) | 5 | Restricted age ranges yielded smaller number of participants (n=1,086) and may further limit representation of younger and older adults. |
|  | **BMI Restrictions** | Included (Y) | 20 | Trials that applied BMI thresholds had lower female participation (41.0%) and tended to have larger sample sizes (mean = 451.1) |
|  |  | Not Included (N) | 7 | Trials without BMI limits enrolled slightly smaller cohorts (mean = 406.0) but had substantially higher female representation (55.5%) |
|  | **Comorbidity Exclusions** | Yes (explicitly stated) | 6 | Excluding based on comorbidities was associated with smaller cohort sizes (mean = 247) and lower female enrolment (43.7%) |
|  |  | No / Not Reported | 21 | A lack of explicit comorbidity exclusion criteria was associated with larger cohorts (mean = 541) and more balanced gender representation (49.8%) |
| **Mental Health** | **Age** | Broad (18–75 years) | 19 | Broader age limits captured most participants (n=6,349); however, enrolled cohorts remained concentrated in early to mid-adulthood, despite eligibility extending into older ages. |
|  |  | Restricted (18–60 years) | 9 | Narrower age bands yielded fewer participants (n=1,812) and further restricted representation of older adults (>60 yrs). |
|  | **Gender / Sex** | Open to all sexes (no sex-based exclusions) | 28 (Male dominance in 64.3% of trials) | Despite universal eligibility, men were more frequently represented, suggesting gender differential enrolment rather than eligibility constraints. |
|  | **BMI Criteria** | BMI restrictions applied | 2 | BMI based criteria rarely used and inconsistently reported; insufficient evidence to determine effects on enrolment. |
|  |  | No BMI restrictions | 26 | Majority of trials placed no BMI constraints, supporting broad physical eligibility but with unclear effects on cohort composition. |
